# Predictors of COVID testing among Australian youth: Insights from the Longitudinal Study of Australian Children

**DOI:** 10.1101/2021.10.31.21265627

**Authors:** Md Irteja Islam, Verity Chadwick, Alexandra Martiniuk

**Affiliations:** Maternal and Child Health Division, International Centre for Diarrheal Disease Research, Bangladesh (icddr,b), 68, Mohakhali, Dhaka 1212, Bangladesh; Sydney School of Public Health, Faculty of Medicine and Health, The University of Sydney, Edward Ford Building, A27 Fisher Road, NSW 2006, Australia; Centre for Health Research and School of Business, The University of Southern Queensland, West Street, Darling Heights, QLD 4350, Australia; Royal North Shore Hospital, Reserve Rd, St Leonard’s NSW 2065, Australia; Office of the Chief Scientist, The George Institute for Global Health, Level 5/1 King Street, Newtown NSW 2042, Australia; Dalla Lana School of Public Health, The University of Toronto, 155 College St Room 500, Toronto, ON M5T 3M7, Canada

**Author notes:** Corresponding author: Md Irteja Islam PhD, Assistant Scientist, Maternal and Child Health Division (Level 7), International Centre for Diarrheal Disease Research, Bangladesh (icddr,b), 68, Mohakhali, Dhaka 1212, Bangladesh.

**Keywords:** COVID-19, SARS-COV-2, Coronavirus, Australia, adolescents, young adults, testing

## Abstract

**Background:** Testing has played a crucial role in reducing the spread of COVID. Although COVID symptoms tend to be less severe in children and adolescents, a key concern is young people’s role in the transmission of the virus given their highly social lifestyles. In this study, we aimed to identify the predictors associated with COVID testing in Australian youth using data from the Longitudinal Study of Australian Children (LSAC).

**Methods:** We used the latest wave 9C1 of the LSAC, where data were collected from 16–21-year-old Australians via an online survey between October and December 2021. In total, 2291 Australian youths responded to the questions about COVID testing and COVID symptom severity. Data was stratified by living with/without parents, and bivariate and logistic regression analyses examined predictor variables (age, sex, country of birth, remoteness, education level, employment, relationship status, number of household members, living with parents, receiving the COVID financial supplement from government and index of relative socio-economic advantage and disadvantage) and their distributions over the outcome variable COVID testing.

**Results:** Youths aged 16-17 were more likely to live at home than youths aged 20-21 years. The strongest predictor of COVID testing was living in major cities (regardless of living with or without parents). Changed household composition was significantly associated with COVID testing among the youths living in the parental home. While among the respondents living without their parents, living with multiple household members and low or no cohesion among household members was associated with higher rates of COVID testing.

**Conclusion:** Our study revealed young people have been very good at getting tested for COVID. To further incentivise testing in this age group, we should consider providing this age group with continued financial and social support while awaiting the outcome of the test and during any isolation.

**Strengths and limitations of this study:**

- Large national cohort of young people strengthened the findings of the study and allowing us to examine the factors associated with COVID testing for the first time in Australia.
- A broad-based assessment of potential predictors of COVID testing, including sociodemographic and coronavirus specific factor.
- Cross-sectional observational design limits causal inference.
- Self-reported information about COVID testing can be subject to recall as well as social desirability bias.

## Introduction

Public health interventions for COVID including testing and isolating, social distancing and bans on mass gatherings, wearing of masks, improving ventilation, closing schools, and working remotely, have been effective worldwide in reducing the spread of the SARS-COV-2 virus.^1^ With the introduction of COVID vaccines, governments, medical and public health experts have been devising exit strategies to resume more ‘pre-COVID’ ways of life. While the vaccines are effective in reducing hospitalisation and mortality rates,^2^ there are some caveats. Evidence demonstrates that vaccinated people can still transmit the virus to others, there is decreased efficacy in the elderly and immunocompromised,^3^ immunity tends to wane over time,^4^ some people have not had access to a vaccine and some people may choose not to get vaccinated.^5^

Testing, tracing, and isolation/quarantine (TTIQ) play crucial roles in epidemic control, which requires fast and comprehensive testing and tracing, and then sufficient compliance with isolation/quarantine.^6^ At this point in time, regular testing is mandatory in some contexts, such as some workplaces or during travel.^7, 8^ However, in other situations we must rely on individuals to recognise the need to be tested and to promptly attend for a test.

It is well established that all people, but more so children and young people, can be asymptomatic carriers of SARS-CoV-2.^9^ Although COVID symptoms and outcomes tend to be less severe in children and adolescents, a key concern is that young people may be important community reservoirs for the transmission of the virus to household members, grandparents, and other community members. Further, adolescents and young adults tend to have a higher number of in-person connections on a daily basis,^10^ and often live in crowded houses^11^ and work multiple jobs.^12^ These factors increasing the likelihood of adolescents and young adults playing a crucial role in SARS-COV-2 transmission as societies open up.^13-15^

Limiting young people from attending education institutions and places of employment cannot be a long-term solution. School and university closures adversely affect young people as they miss out on education (particularly those from disadvantaged backgrounds),^16^ opportunities to develop social skills^17^ and find role models from outside the home.^18^ Indeed, 30% of Australians aged 18 to 34 years experienced high or very high levels of psychosocial distress in June 2021, almost double the rate reported by this age group in 2018-19.^19^ Distress was greatest in Melbourne and Sydney where the tighter lockdowns occurred, while the prevalence of high or very high psychosocial distress in other age groups remained relatively unchanged when compared to the year prior to COVID.^19^

Throughout the whole pandemic period to date, just under half of Australians reported they would definitely get a COVID test if experiencing mild respiratory symptoms.^20, 21^ NSW statistics suggest that approximately one quarter of daily COVID tests have been for individuals aged 10-29 years of age.^22^ Although testing is crucial for all age groups, testing is arguably more vital in the highly social and mobile adolescent and young adult age group.^10^ However, there is little knowledge regarding predictors of adolescents and young adults seeking COVID testing. Here we examine what factors are associated with rates of COVID testing in 16–21-year-old Australians.

## Methods

This paper is being reported according to STROBE guidelines for observational studies. ^23^ Study design and Sample

We used data from Growing Up in Australia: The Longitudinal Study of Australian Children (LSAC), a population-based cross-sequential cohort study carried out by the Australian Government Department of Social Services (DSS) in a partnership with the Australian Institute of Family Studies (AIFS) and the Australian Bureau of Statistics (ABS). The LSAC sampled participants from the Medicare enrolment database, Australia’s universal health insurance scheme. A multi-stage cluster sampling design was used. First, representative postcodes were selected employing probability proportion to size method, stratified by state or territory and by capital city statistical division vs. rest of state to guarantee geographically proportionate samples across urban and rural areas. Second, children were randomly selected from a selection of 311 postcodes, around 40 and 20 children per postcode in the large and small states, respectively.^24^

The LSAC collected data biennially since 2004 from two cohorts - the birth cohort (B cohort) aged 0–1 year at baseline (n=5107) and the kindergarten cohort (K cohort) aged 4–5 years at baseline (n=4983). Out of the total 10090 children recruited during Wave 1, 2958 responded in Wave 9C1 (i.e., the latest online survey data collected during COVID pandemic in between October-November 2020 among 16-17-year-olds B cohort and 20-21-year-olds K cohort). In the LSAC, data were collected primarily from the child’s parents (typically from the mother or Parent 1) with additional information from other caregivers (such as Parent 2, usually the father) and the children themselves (when aged 10 years or more for some specific questions). Data collection methods for the prior Waves 1-8 included face-to-face interview, mail-out questionnaires, time-use diaries, Computer-Assisted Self-Interview (CASI) and Computer-Assisted Web-Interview (CAWI); while due to COVID pandemic in 2020, only CAWI was used to collect data from parents and youth in LSAC Wave 9C1. More details on the LSAC methodology, including sampling procedures and data collection techniques, is described elsewhere.^24, 25^

Our current study included 2291 Australian youth aged 16-21 years at the time of the LSAC Wave 9C1 in 2020. Out of them, 1374 (60%) and 917 (40%) youth were from B-cohort and K-cohort, respectively. Respondents who reported on the outcome variable (COVID testing) and explanatory variables were included in the analyses; while the ‘Missing’, ‘Don’t know’, ‘Prefer not to say’ and ‘Refused’ responses categories were omitted (n=667).

### Measures

The LSAC Wave 9C1 online survey questionnaire included questions related to the COVID pandemic (including the coronavirus restriction period between March-May 2020) and sociodemographic characteristics among the parents and/or youths.

### Outcome variable

The question related to testing for COVID included whether the youth have been tested for severe acute respiratory syndrome coronavirus 2 (SARS-CoV-2) with response categories ‘Yes’ (coded as 1) and ‘No’ (coded as 0).

### Explanatory variables

Based on the previous literature,^26, 27^ the following socio-demographic variables were considered as predictor variables in this study: age (16-17 years, 20-21 years), sex (Male, Female), country of birth (Australia, Overseas), remoteness (Major cities, Rural/remote areas), education (Technical/others, Secondary, University/tertiary), employment (Full-time, Part-time, Unemployed), number of household members (Alone, Two people, Three people, Four or more people), living with parents (Yes, No), currently in a relationship (Yes, No), and the SEIFA (Socio-Economic Indexes for Areas) Index of Relative Socio-Economic Advantage and Disadvantage (IRSAD) quintiles. For all areas across Australia, IRSAD quintile 1 includes the lowest 20 percent for the most disadvantaged areas, and quintile 5 contains the highest 20 percent for the most advantaged areas ^28^. Regarding family cohesion, in general, the youth were directly asked to rate the ability of family or household members to get along with each other where the responses were recorded in a 5-point scale (from Excellent to Poor).^29^ However, for analytical purposes, we created a dichotomized variable – ‘cohesion among household members’ from the responses. Youth who responded ‘excellent’, ‘very good’ or ‘good’ were classified as ‘yes’ (coded as 0), while those who answered, ‘fair’ or ‘poor’ were classified as ‘no’ (coded as 1).

Since, the respondents of LSAC Wave 9C1 experienced the COVID pandemic in 2020, the study included COVID and coronavirus restriction period (CRP) variables between March-May 2020 in Australia.^25^ Our study thus included the following additional variables: employment status during CRP (Employed, Unemployed), changed household composition during CRP (Yes, No), and received coronavirus supplement^1^ during CRP (Yes, No) to assess whether these variables are associated with COVID testing among the respondents. The following question was directly asked to youth in W9C1 about the changes in household composition since the previous LSAC W8 in 2018 as the data were rolled forward: ‘Whether the household composition changed during COVID-19 restrictions?’. Note that the regular detailed household data could not be gathered in W9C1 since the questions were meant to be asked in a face-to-face interview (which was not possible during CRP) and could not be fully converted into online inquiries.^25^

### Statistical analysis

To understand the sociodemographic predictors of COVID testing among youths, stratified by living with/without parents, initially, we computed summary statistics. Then, we conducted bivariate analyses to examine the predictor variables and their distributions over the outcome variables (COVID testing). Pearson’s chi-squared test^31^ signified the strength of the bivariate associations between the characteristics and youth COVID testing, stratified by living with/without parents. Further, we employed logistic regression analysis to identify predictors of COVID testing. The results of the regression analyses were presented as adjusted odds ratios (aOR) with corresponding 95% confidence intervals (CI) after adjusting for possible confounding effects of other explanatory variables. All statistical analyses were conducted with Stata/MP 14.1, and ‘svy’ command^32^ was used to take account for of the LSAC’s multi-stage clustered sampling design, and to deal with potential bias and to avoid overestimation of the significance.

### Patient and public involvement

The LSAC is a population-based longitudinal study, hence no patient groups were engaged in its design or execution. To the best of our knowledge, the public was not involved in designing, recruiting, or implementation of the LSAC study. Parents received a brief health report for their children and themselves during or shortly after the assessment visit. They agreed to participate understanding that they would not otherwise get individual results about themselves or their children.

### Ethics

The LSAC has been approved by the Human Research Ethics Committee of the Australian Institute of Family Studies (AIFS) (Application number 20-09), and written informed consent was obtained for all study participants. In addition, the authorship team obtained permission from Australian Data Archive Dataverse to use LSAC data for research and publications (Reference No. 263493).

## Results

Descriptive statistics of the study population are demonstrated in Table 1. Of the 2291 youth, 60% were aged between 16-17 years, more than 56% were female, 97% were born in Australia, 73% were from major cities, 57% had secondary level educational qualifications, and 57% had part-time employment. About 75% of youth had three or more household members, 84% reported cohesion among the household members, 88% of youth were living with parents and 30% were in a relationship. Nearly 70% of respondents were from the disadvantaged group (Quintile 1, Quintile 2 and Quintile 3 combined). During the coronavirus restriction period (CRP) between March-May 2020, more than 50% of youth were unemployed, the majority of the respondents (84%) did not experience changed household composition, and 82% of youth did not receive the coronavirus financial supplement.

**Table 1.**
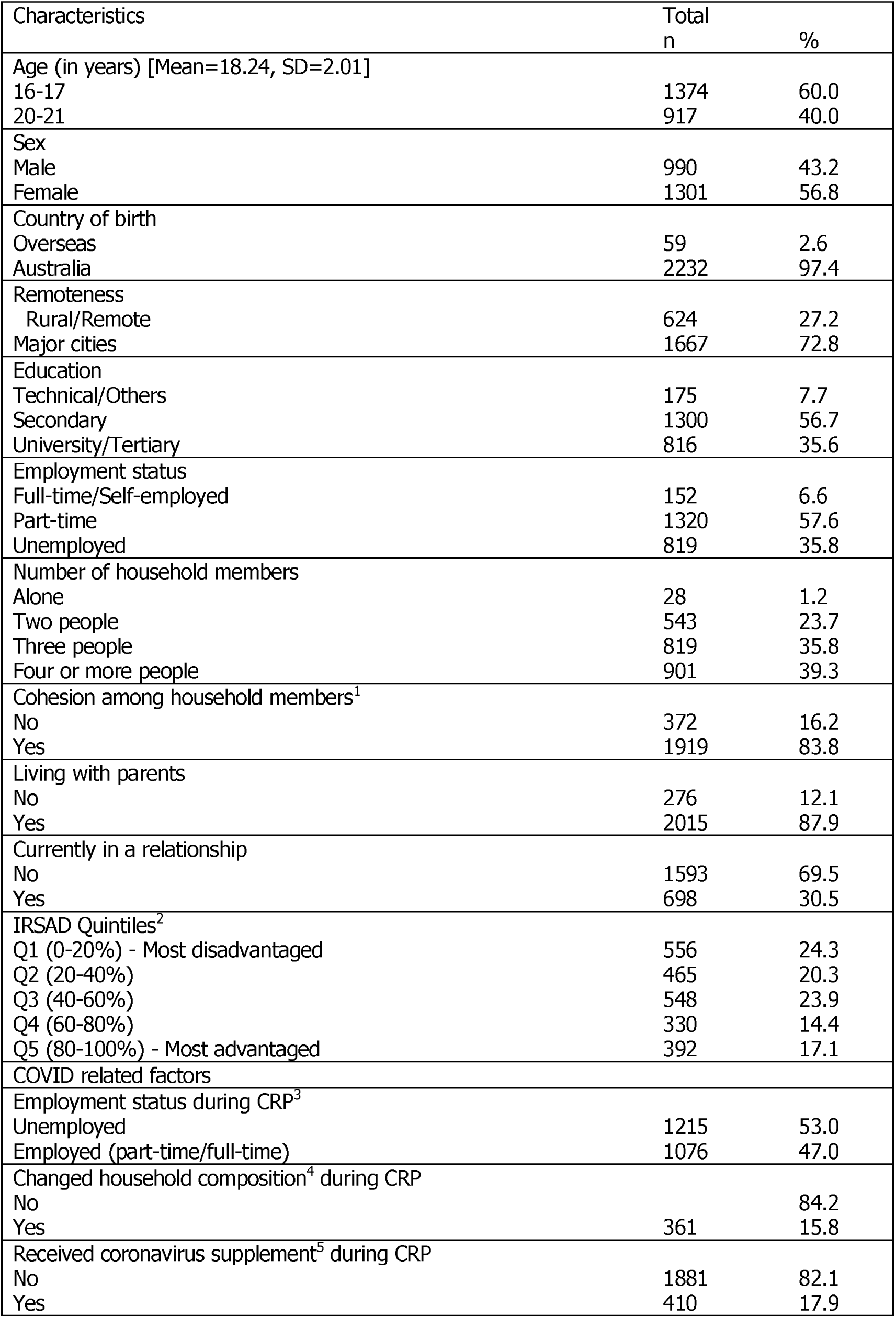
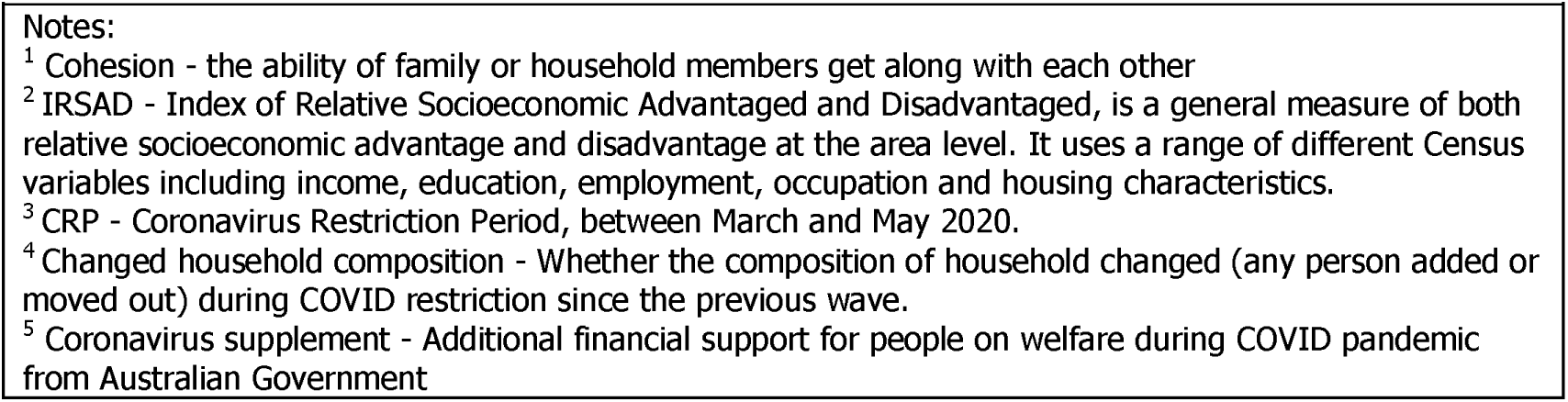
Sample characteristics (n=2291)

Fig. 1 displays the percentages of youth tested for COVID, stratified by living status. Overall, 587 (26%) youth got tested for COVID. Out of which, 85% of youth were living with parents, and 15% were living without parents.

**Figure 1.**
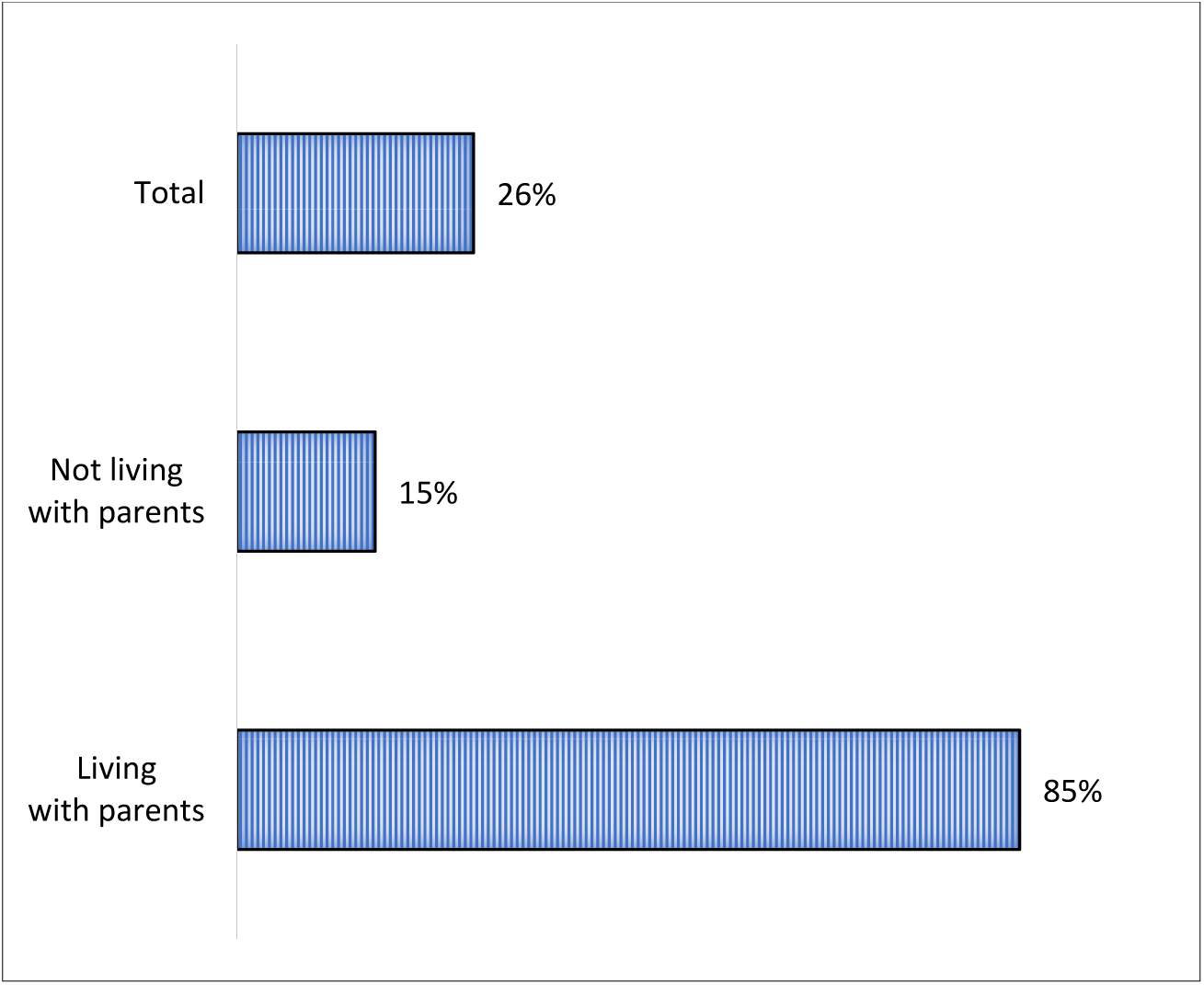
Percentages of youth been COVID tested, stratified by living with and without parents

Bivariate relationships between explanatory variables and COVID testing among youths, stratified by living status are reported in Table 2. Among those who were living with their parents, more than 60% (p=0.009) of youth were those aged 16-17 years compared to the age group 20-21 years. Youth who were living in major cities compared to those living in rural/remote areas, and youth with changed household composition during CRP than their counterparts were also found to be factors associated with COVID testing among the young people who were living with their parents (p<0.010 for all). While for youths not living with parents, Table 2 also shows that remoteness and cohesion among household members were significantly associated with COVID testing (p<0.05 for all).

**Table 2.**
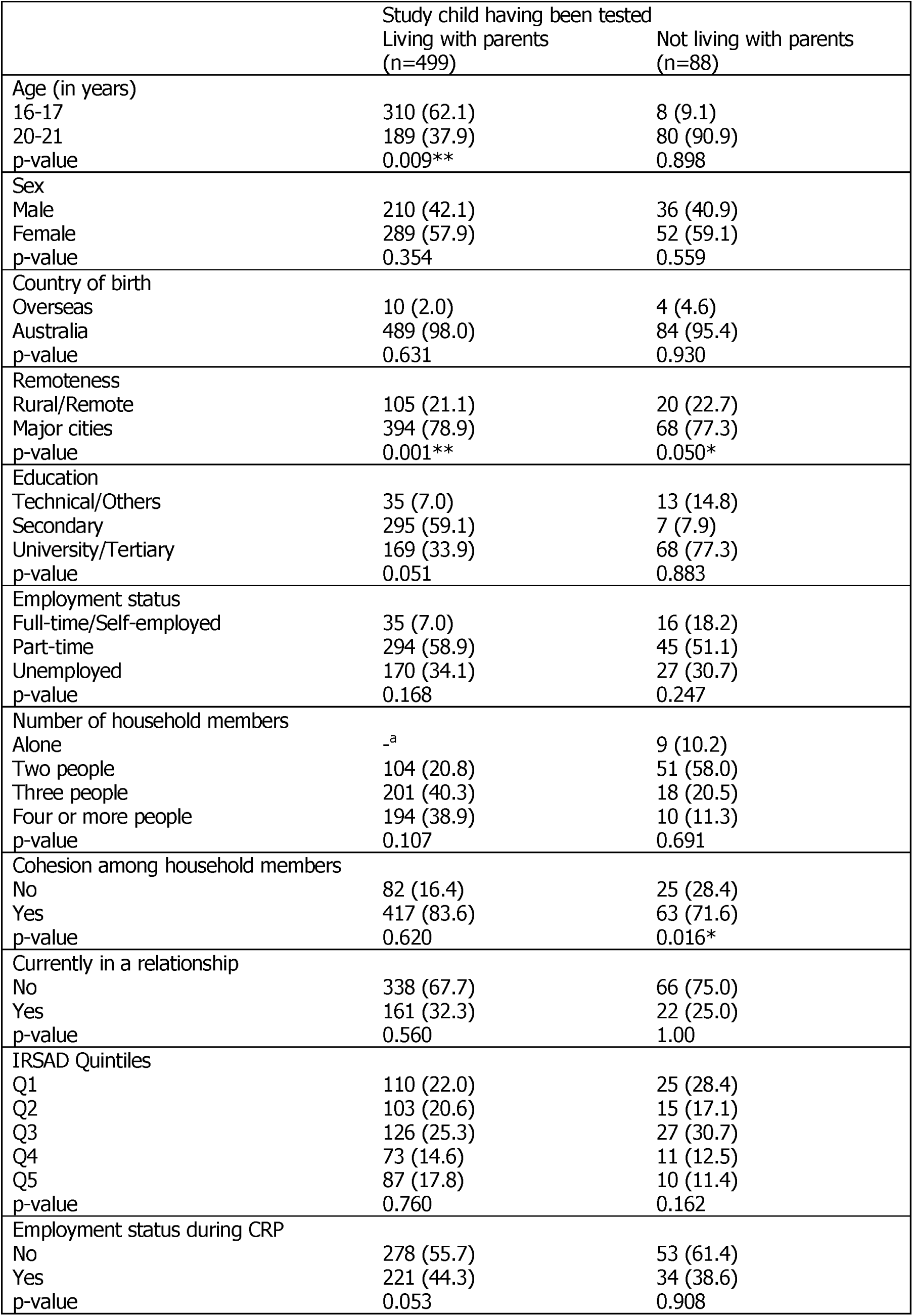
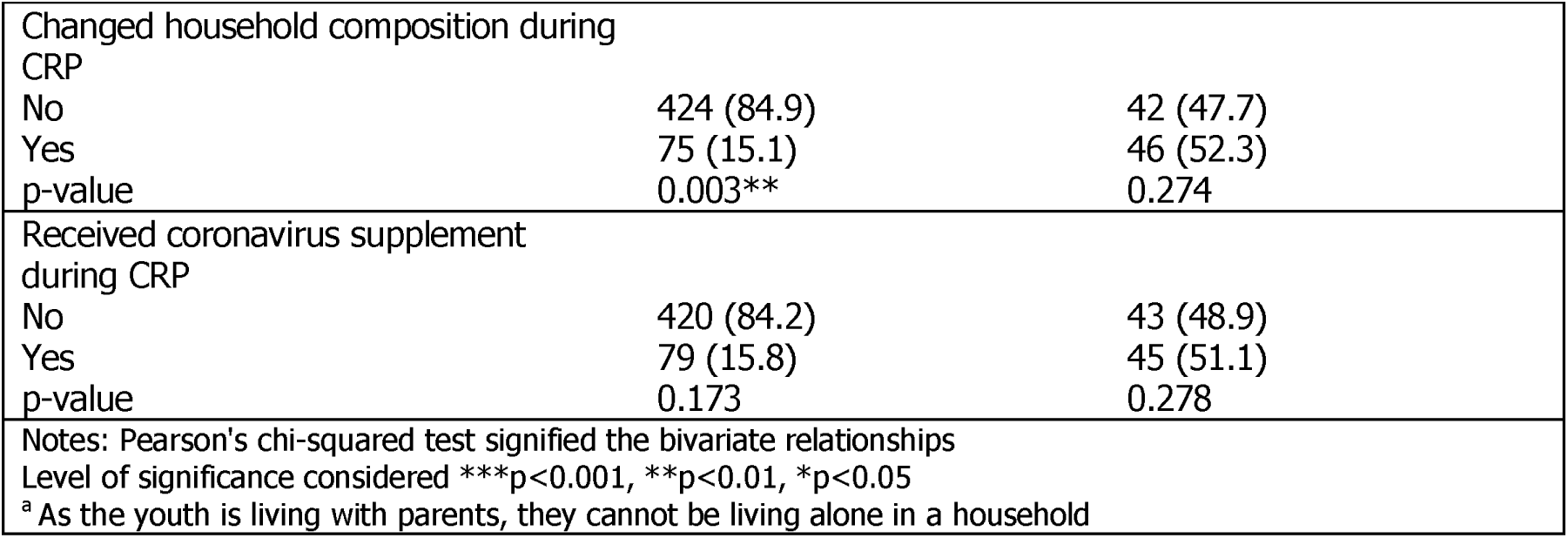
Bivariate associations between factors and COVID testing, stratified by youth living with/without parents

|  | Study child having been tested |  |
| --- | --- | --- |
|  | Living with parents<br>(n=499) | Not living with parents<br>(n=88) |
| Age (in years) |  |  |
| 16-17 | 310 (62.1) | 8 (9.1) |
| 20-21 | 189 (37.9) | 80 (90.9) |
| p-value | 0.009** | 0.898 |
| Sex |  |  |
| Male | 210 (42.1) | 36 (40.9) |
| Female | 289 (57.9) | 52 (59.1) |
| p-value | 0.354 | 0.559 |
| Country of birth |  |  |
| Overseas | 10 (2.0) | 4 (4.6) |
| Australia | 489 (98.0) | 84 (95.4) |
| p-value | 0.631 | 0.930 |
| Remoteness |  |  |
| Rural/Remote | 105 (21.1) | 20 (22.7) |
| Major cities | 394 (78.9) | 68 (77.3) |
| p-value | 0.001** | 0.050* |
| Education |  |  |
| Technical/Others | 35 (7.0) | 13 (14.8) |
| Secondary | 295 (59.1) | 7 (7.9) |
| University/Tertiary | 169 (33.9) | 68 (77.3) |
| p-value | 0.051 | 0.883 |
| Employment status |  |  |
| Full-time/Self-employed | 35 (7.0) | 16 (18.2) |
| Part-time | 294 (58.9) | 45 (51.1) |
| Unemployed | 170 (34.1) | 27 (30.7) |
| p-value | 0.168 | 0.247 |
| Number of household members |  |  |
| Alone | _a | 9 (10.2) |
| Two people | 104 (20.8) | 51 (58.0) |
| Three people | 201 (40.3) | 18 (20.5) |
| Four or more people | 194 (38.9) | 10 (11.3) |
| p-value | 0.107 | 0.691 |
| Cohesion among household members |  |  |
| No | 82 (16.4) | 25 (28.4) |
| Yes | 417 (83.6) | 63 (71.6) |
| p-value | 0.620 | 0.016* |
| Currently in a relationship |  |  |
| No | 338 (67.7) | 66 (75.0) |
| Yes | 161 (32.3) | 22 (25.0) |
| p-value | 0.560 | 1.00 |
| IRSAD Quintiles |  |  |
| Q1 | 110 (22.0) | 25 (28.4) |
| Q2 | 103 (20.6) | 15 (17.1) |
| Q3 | 126 (25.3) | 27 (30.7) |
| Q4 | 73 (14.6) | 11 (12.5) |
| Q5 | 87 (17.8) | 10 (11.4) |
| p-value | 0.760 | 0.162 |
| Employment status during CRP |  |  |
| No | 278 (55.7) | 53 (61.4) |
| Yes | 221 (44.3) | 34 (38.6) |
| p-value | 0.053 | 0.908 |
|  | Living with parents<br>(n=499) | Not living with parents<br>(n=88) |
| Changed household composition during<br>CRP |  |  |
| No | 424 (84.9) | 42 (47.7) |
| Yes | 75 (15.1) | 46 (52.3) |
| p-value | 0.003** | 0.274 |
| Received coronavirus supplement<br>during CRP |  |  |
| No | 420 (84.2) | 43 (48.9) |
| Yes | 79 (15.8) | 45 (51.1) |
| p-value | 0.173 | 0.278 |
| Notes: Pearson's chi-squared test signified the bivariate relationships<br>Level of significance considered ***p<0.001, **p<0.01, *p<0.05<br><sup>a</sup> As the youth is living with parents, they cannot be living alone in a household |  |  |

Results from logistic regression models of respondents being tested for SARS-CoV-2 testing are portrayed in Table 3. The strongest predictor of COVID testing was living in major cities regardless of whether the young person was living with parents (aOR 1.82, 95% CI: 1.27-2.60) or without parents (aOR 2.25, 95% CI: 1.20-4.20) compared to those who were living in rural/remote areas. Among those who were living with parents, changed household composition during the coronavirus restriction period (CRP) increased the probability of COVID testing (aOR 1.33, 95% CI: 0.98-1.81) compared to their counterparts. Among the respondents who were living without parents, living with multiple household members (Two people: aOR 5.92, p<0.001; three people: aOR 5.34, p<0.001; Four or more people: aOR 2.66, p<0.05) significantly increases the likelihood of COVID testing compared to those who lived alone. Moreover, no cohesion among the household members (aOR 6.03, 95% CI: 2.40-15.16) increases the probability of getting COVID tested among the youth who were not living with parents compared to those who had a cohesive household.

**Table 3.** Logistic regression models for youth having been tested

|  |  | Youth having been tested<br>Living with parents |  | Not living with parents |  |
| --- | --- | --- | --- | --- | --- |
|  |  | Adjusted OR | 95% CI | Adjusted OR | 95% CI |
| Age (Ref. 16-17) | 20-21 | 1.54 | 0.75-3.16 | 2.75 | 0.29-25.75 |
| Sex (Ref. Male) | Female | 1.17 | 0.94-1.46 | 1.29 | 0.55-3.01 |
| Country of birth (Ref. Overseas) | Australia | 1.05 | 0.37-2.98 | 1.64 | 0.31-8.47 |
| Remoteness (Ref. Rural/Remote) | Major cities | 1.82** | 1.27-2.60 | 2.25* | 1.20-4.20 |
| Education (Ref. Technical/Others) | Secondary | 1.41 | 0.68-2.89 | 2.50 | 0.18-33.84 |
|  | University/Tertiary | 0.92 | 0.62-1.39 | 0.60 | 0.18-1.95 |
| Employment status (Ref. Full-time/Self-employed) | Part-time | 0.78 | 0.44-1.39 | 0.53 | 0.20-1.39 |
|  | Unemployed | 0.84 | 0.46-1.55 | 0.74 | 0.29-1.89 |
| Number of household member (Ref. Lived alone) | Two people | -.<sup>a</sup> | - | 5.92*** | 2.48-14.13 |
|  | Three people | 1.05 | 1.94-5.92 | 5.34*** | 2.67-10.66 |
|  | Four or more people | 0.84 | 1.83-9.09 | 2.66* | 1.13-6.27 |
| Cohesion among household members (Ref. Yes) | No | 1.12 | 0.82-1.34 | 6.03** | 2.40-15.16 |
| Currently in a relationship (Ref. No) | Yes | 1.06 | 0.84-1.46 | 1.13 | 0.56-2.29 |
| IRSAD Quintile (Ref. Q1) | Q2 | 0.85 | 0.51-1.40 | 0.51 | 0.15-1.70 |
|  | Q3 | 0.94 | 0.66-1.36 | 0.87 | 0.27-2.76 |
|  | Q4 | 0.82 | 0.60-1.13 | 0.31** | 0.14-0.66 |
|  | Q5 | 0.69 | 0.44-1.09 | 0.62 | 0.26-1.48 |
| Employment status during CRP (Ref. Unemployed) | Employed | 0.81 | 0.57-1.07 | 0.87 | 0.41-1.84 |
| Changed household composition during CRP (Ref. No) | Yes | 1.33* | 0.98-1.81 | 1.29 | 0.52-3.16 |
| Received coronavirus supplement during CRP (Ref. No) | Yes | 1.04 | 0.78-1.31 | 0.85 | 0.35-2.04 |
| Level of significance: ***p<0.001, **p<0.01, *p<0.05 |  |  |  |  |  |
| <sup>a</sup> For youth living with parents, 'Two people' used as Ref category since they cannot be living alone in a household |  |  |  |  |  |

## Discussion

This study examined factors associated with COVID testing in adolescents and young adults. Those living with their parents were more likely to get a COVID test, and this was especially true if there had been a change in their household composition. Among those not living with their parents, youths were more likely to get a COVID test if they were living in major cities, if they had non-cohesive households, and if they were in households consisting of two or more people.

Higher COVID testing rates were observed for adolescents and young adults who were living with their parents. This may be related to parents monitoring their children for symtpoms of COVID, and encouraging and taking them to testing facilities if symptoms were present.^33^ Other reports have found parents are more likely to have their children tested if other family members lived at home, or if the family was in continual contact with relatives or at-risk individuals.^34^ This could relate to parents seeking COVID testing to protect others - ultimately pro-social behaviour.^33^

Changes in family composition were also associated with higher test rates when adolescents lived at home. For example, this could be attributed to family members moving to live with grandparents, parents separating, or children moving back home. This may be associated with a larger number of family members at home and therefore an increased risk of COVID entering the household.^34^ It could also be related to family members in different homes wanting returning household members (for example, children going between families) to get tested to protect household members.^35^ Previous research during the COVID pandemic has shown that public health restrictions have led to negative impacts on spousal relationships, ^36^ with higher rates of domestic violence^37^ and divorce compared to pre-pandemic.^38^ We were not able to capture the extent to which household composition changes were related to separating parents within the coronavirus restriction period, but the Family Court of Australia did report a 39% increase in parenting-related disputes in the second month of the pandemic.^39^ This may be attributed to parents in different homes expressing concern about the other household’s adherence to social distancing and mask wearing,^35^ or the number of people in contact with other households as reported elsewhere.^35, 40^

Further, houeholds with low or no cohesiveness (a measure of connectivity and caring within households) increased the testing rates for young people living outside of parental home. Low cohesiveness could mean that these youths cannot trust their household members to adequately protect themselves from COVID. In turn, their higher testing rates may be an effort to protect their families when visiting the family home, supported by parents who are mindful of the risks of COVID and recommend protective measures.^41^ Government promotions have framed the motivation as helping the community rather than avoiding individual risk,^42^ or using identity-based messages such as “don’t be a spreader”.^43^ However, those living out of home with more secure financial footing were the least likely to have COVID tests, as indicated by the significant negative relationship between quintile 4 IRSAD and COVID testing. This could reflect possible instability in their employment position and unwillingness to risk calling in sick to isolate if testing positive.

For adolescents not living at home with their parents, factors associated with increased testing rates included living in urban areas, and the likelihood of testing increased with presence of multiple houshold members. Urban areas have experienced greater outbreaks of COVID^44^ given increased living density, and connectivity to people.^45^ In Australia, this is also because most cases (prior to June 2021) were imported from overseas and spread into the urban community from quarantine facilities based in cities. Furthermore, people living outside of urban areas may have less access to testing. Recent studies have found that the one of the most common reasons for symptomatic individuals not getting tested was not knowing where to go.^46, 47^ COVID testing rates for adolescents were lower in rural communities, and more should be done to increase these rates. Given the common combination of poverty, the higher prevalence of comorbid diseases, and poor accessibility to healthcare,^48^ individuals in rural settings are likely to experience worse outcomes when COVID outbreaks occur, as was the case for the Wilcannia outbreak in rural New South Wales.^49^

The majority (70%) of our study respondents were socioeconomically disadvantaged, but despite this over a quarter were tested for COVID. In the future, this age group may be dissuaded from getting tested given they may be vaccinated, asymptomatic or only mildly affected, and they may not want to isolate after testing given the potential loss of casual-work income and socialising opportunity. Some studies of predictors of COVID testing have been published in other populations. In the USA, factors predicting testing included: free testing, concern about health, and having a high number of social contacts.^10, 50^ Interestingly, in a study using hypothetical scenarios, individuals said cost and ability to support oneself if needing to isolate were of no concern and did not influence their likelihood to get tested.^10^ However, in practice American Latino and Black respondents and those experiencing financial strain were disproportionately likely to indicate that resource factors would influence their decision to get tested.^50^ This effect remained after controlling for being uninsured and experiencing material hardship and general financial constraints.^50^ Such findings are consistent with other research^51, 52^ indicating that basic survival needs of individuals and families often outweigh other consideration in healthcare decision-making, including prosocial motives like avoiding transmission of infection to others. Further studies demonstrate that the reasons adolescents do not participate in asymptomatic COVID testing include concerns about the mental health impact of self-isolation, the impact on others if the test is positive (such as forcing others into isolation),^53^ and thinking that it is a potential waste of resources.^54^ More effective economic support for youth may assist in supporting this age group to be tested for COVID.

The generalisability of these findings may be limited to Australian young people given that the sample was not representative of the Australian overall population. Another limitation may be the difficulty in comparing these findings to other countries, for example, Australia’s COVID testing rate is much higher than many other countries; 2.7 per day per 1,000 people^55^ compared to the US (1.9 per 1,000), the UK (1.3 per 1,000) and India (0.654 per 1,000).^56^ This may be due to Australia’s relatively high national wealth (and lower inequality compared to US), as well as finanical support for individuals and businesses afflicted by the pandemic.^57^

## Conclusion

In conclusion, adolescents and young adults living with their parents were more likely to be tested for COVID compared to those who lived outside of the family home, and changes in who lived at home during the pandemic was associated with higher testing rates. For youths not living at home, living in the city, living with two or more housemates, and for participants reporting poor unity of households, testing rates were higher. In conclusion, while young people have been very good at getting tested for COVID, to further incentivise testing in this age group, we should consider providing financial and social support while awaiting the outcome of the test and during any isolation.

## Data Availability

The LSAC dataset is publicly available from the National Centre for Longitudinal Data (NCLD) Dataverse on request available at https://growingupinaustralia.gov.au/data-and-documentation/accessing-lsac-data.

## Data availability statement

The LSAC dataset is publicly available from the National Centre for Longitudinal Data (NCLD) Dataverse on request available at https://growingupinaustralia.gov.au/data-and-documentation/accessing-lsac-data. Authors do not have permission to share this unit record data without endorsement from the Department of Social Services and Australian Institute of Family Studies.

## Ethics statements

Patient consent for publication

Not applicable.

Ethics approval and consent to participate

The LSAC has been approved by the Human Research Ethics Committee of the AIFS, and written informed consent was obtained for all study participants. In addition, the authorship team obtained permission from Australian Data Archive Dataverse to use LSAC data for research and publications.

## Author’s contribution

MII conceptualized the study, analyzed the data, drafted parts of the initial manuscript, reviewed and revised the manuscript. VC drafted parts of the initial manuscript, and reviewed and revised the manuscript. AM conceptualized the study, drafted parts of the initial manuscript, reviewed and revised the manuscript. All authors approved the final manuscript as submitted and agree to be accountable for all aspects of the work.

## Footnotes

1 An additional top-up payment for people on welfare during COVID-19 pandemic in Australia 30. Australia S. Coronavirus Supplement, https://www.servicesaustralia.gov.au/individuals/services/centrelink/coronavirus-supplement (2021, accessed 19 October 2021 2021).

